# Usability, acceptability and feasibility of continuous glucose monitoring among children and adolescents with type 1 diabetes in Kenya

**DOI:** 10.64898/2026.08.27.26361447

**Authors:** Prisca Amolo, Lucy Mungai, Agnes Karingo Karume, James Kibugi, Winfred Mwende, Núria Botella, Cathy Haldane, Yvonne Kamau, Elena Marbán-Castro

## Abstract

**Introduction:** Continuous Glucose Monitoring (CGM) is considered standard care in high-income countries. There is, however, limited published evidence on CGM use in low– and middle-income countries. The purpose of this study was to assess the usability, acceptability, and feasibility of CGM use among people living with type 1 diabetes (T1D) and caregivers in a low-resource setting.

**Research Design and Methods:** This prospective study conducted at the Kenyatta National Hospital purposively enrolled persons aged 4-25 years who had been on management for T1D for at least six months, and caregivers of those under 18 years. Fourty youth living with T1D used CGM for three months in place of self monitoring of blood glucose (SMBG). The System Usability Scale (SUS), a Theoretical Framework of Acceptability-based questionnaire, the Diabetes Distress Scale (DDS), the Glucose Monitoring Satisfaction Survey (GMSS), and a feasibility survey were administered. Outcomes were summarized descriptively, including means, medians, and frequencies using R statistical software.

**Results:** The median SUS score was 98.8 (IQR 92.5-100.0). Acceptability was high, and the median total GMSS score improved from 3.73 to 4.73. Among adolescents and adults, the median overall DDS score reduced from 1.54 to 1.36, with reductions in scores in all domains, except for “hypoglycemia distress” which increased, and “physician distress” which remained low. Among caregivers, the median overall DDS score declined from 2.05 (moderate distress) to 1.90 (low distress), with modest reductions in “teen management” and “parent-teen relationship distress” and a slight increase in “personal distress”. Median CGM active wear time was 89%.

**Conclusion:** This study comprehensively evaluated CGM across usability, acceptability, and feasibility outcomes, with the findings supporting the integration of CGM into routine diabetes management in low-resource settings. The short follow-up period, however, may not capture changing perceptions or long-term adherence.

**What is already known on this topic:**

- Self-monitoring of blood glucose is integral to type 1 diabetes (T1D) management but is associated with pain, inconvenience, and limited ability to capture fluctuations in glucose levels. Continuous glucose monitoring (CGM) offers a solution by providing real-time, minimally invasive glucose measurements, thereby facilitating better diabetes care. However, in low– and middle-income countries CGM is not routinely available, and data on its usability, feasibility, and acceptability remain limited.

**What this study adds:**

- There was high usability, acceptability, and feasibility of CGM among youth with T1D, with sustained engagement, improved glucose monitoring satisfaction, and modest reductions in diabetes distress.

**How this study might affect research, practice or policy:**

- This study’s findings support the wider adoption of CGM for management of youth with T1D in resource-limited settings.

## Introduction

Type 1 Diabetes mellitus (T1D) is a chronic autoimmune condition characterized by persistent hyperglycemia due to absolute insulin deficiency and is the commonest form of diabetes in the young.^1,2^ Effective management requires lifelong insulin therapy alongside regular blood glucose monitoring.^3^ The Diabetes Control and Complications Trial (DCCT) demonstrated that intensive glycemic management significantly delays the onset and progression of microvascular complications.^4^ This requires either multiple daily injections (MDI) or continuous subcutaneous insulin infusion (CSII), accompanied by frequent self-monitoring of blood glucose (SMBG).^5,6^ Traditionally, SMBG is performed through capillary finger-prick sampling using glucometers. However, this method may fail to detect hypo– or hyperglycemic episodes occurring between tests and is associated with pain, inconvenience, and needle-related anxiety, especially in newly diagnosed children.^3^

Over the past two decades, advances in technology have led to the development and widespread adoption of Continuous Glucose Monitoring (CGM) systems, now considered standard of care in high-income countries.^8^ CGM devices measure glucose concentrations in interstitial fluid, via a subcutaneous sensor typically inserted in the arm or abdomen.^8^ The sensor updates the glucose readings every minute, storing up to eight hours of data, and transmits the data to a reader, receiver, or smartphone application.^9–13^ The International Society for Paediatric and Adolescent Diabetes (ISPAD) recommends continuous use of CGM for all children, adolescents, and young adults living with T1D.^14^ CGM enables identification of glycemic trends and patterns, allowing for more informed decisions around insulin dosing, dietary adjustments, and physical activity.

Much of the available CGM research has been conducted in high-income countries. A recent scoping review on CGM use in low– and middle-income countries (LMICs) identified limited published evidence. Among 27 studies conducted in LMICs, CGM demonstrated positive effects on surrogate outcomes (e.g., HbA1c reduction), hard clinical endpoints (e.g., fewer hospitalizations), and patient-reported outcomes (e.g., quality of life).^15,16^ Nonetheless, the review identified major gaps in scope and volume, underscoring the need for further empirical research, including implementation studies, to inform evidence-based policies in LMICs.

There is a scarcity of research assessing the feasibility and acceptability of CGM in low– resource settings.^17–20^ A 2019 East African pilot and feasibility study, in which 68 participants wore blinded CGM for 2 weeks, found that the sensors were well tolerated, with no adverse events related to the sensor reported.^21^ The aim of this study was to assess the usability, feasibility and acceptability of CGM worn over three months by persons living with T1D and their caregivers, at the Kenyatta National Hospital (KNH) paediatric and adolescent endocrinology clinic.

## Methods

### Study design

This was a pragmatic prospective study; Access to CGMs for Equity in Diabetes Management – Usability (ACCEDE-U), where participants were recruited from routine diabetes clinic visits during which quantitative and qualitative data were collected on usability, acceptability, and feasibility of CGM. Qualitative data is reported separately on a different manuscript.

### Study population

Inclusion criteria were persons living with T1D, aged 4-25 years and who had been on management for T1D for at least 6 months since diagnosis. Parents or caregivers of those aged below 18 years were also included. Exclusion criteria were parents or guardians who were unable to understand how to take care of CGM devices, as indicated by inability to explain back to the researcher.

### Study device

FreeStyle Libre is a CE-approved CGM device which is marketed and available for use in high-income and low-income countries. FreeStyle Libre 2 device was used because unlike other CGM devices, it does not require fingerstick calibrations, making it more user-friendly and reducing participant burden. It provides optional real-time alerts for high and low glucose levels, which can be critical for monitoring participants in real-time. It is also approved for people aged four years and above, making it ideal for this study population. Finally, it is generally more affordable than other CGM devices which is advantageous for research studies and for potential future patient cost.

### Procedures

The study was carried out between November 2025 and March 2026 after obtaining ethical and regulatory approval. KNH is a national tertiary referral hospital in Kenya, and a teaching institution for the University of Nairobi which provides post-graduate and fellowship programs.

The sample size was determined based on the primary objective which was to assess the usability of CGM from the perspective of recipients of care. This sample size was considered sufficient to provide reliable usability assessments using the System Usability Scale (SUS), a validated tool for evaluating user experience. Furthermore, this sample size aligns with best practices in usability research, where studies with 30-50 participants are typically considered adequate to detect meaningful usability trends. A total of 40 participants were recruited, comprising an initial sample of 35 individuals, with an additional 15% (5 participants) included to account for potential attrition. ^22^ Proportional quota sampling was used to select patients based on eligibility criteria, ensuring representation of clinic patients across age groups as follows: 4-9 years (11 patients), 10-15 years (15 patients), 16-20 years (8 patients), and 21-25 years (6 patients).

The study consisted of five visits, including three standard clinic visits, two additional study-related visits (online supplemental table 1). At baseline, after obtaining informed written consent and assent, participants were assisted to complete questionnaires including two standardized surveys Diabetes Distress Scale (DDS), and Glucose Monitoring Satisfaction Survey (GMSS). Information was obtained on the socioeconomic status, diabetes duration, usual insulin regimen, and any recalled episodes of hypoglycemia and hospital admission in the previous three months.

During the second visit participants were educated about the Freestyle Libre CGM system and guided on the application process, and the appropriate measures for maintaining the sensor. On the third visit, which was 14 days after the second visit, the attending physician reviewed downloaded CGM data and provided clinical feedback, including potential insulin regimen adjustments to the participant. Participants returned approximately six weeks later for clinical review and feedback on CGM data, and for qualitative data collection (data reported in a separate manuscript) ^23^.

Finally, at the endline visit, three months after enrollment, the attending physician provided the participant with feedback based on their CGM data, and a follow-up diabetes education session was provided based on each participant’s CGM data insights. Additional standard surveys were administered, including the SUS, the Acceptability survey developed by Sekhon et al, the DDS, GMSS, and a feasibility survey on CGM use.

CGM usability was assessed using the SUS, which is a standardized, ten-item <u>Likert</u> <u>scale</u> used to measure the perceived usability of a system or product. It gives a score from 0 to 100, where higher scores indicate better usability.^24^ Acceptability of CGM was assessed using a structured questionnaire informed by the Theoretical Framework of Acceptability (TFA) by Sekhon et al, which conceptualises acceptability as a multidimensional construct relevant during intervention implementation.^25^

The Glucose Monitoring Satisfaction Survey (GMSS) assessed satisfaction with glucose monitoring modalities, including perceived convenience, intrusiveness, trust in readings, and behavioural burden associated with device use. Items are scored on Likert-type response scales, with higher scores indicating greater satisfaction with monitoring practices. Domain-level scores were derived following published scoring algorithms, allowing comparison across monitoring strategies and timepoints. The instrument has been used in diabetes technology evaluations and captures both experiential and practical dimensions of glucose monitoring.^26^

The Diabetes Distress Scale (DDS) was measured using a validated instrument designed to quantify regimen-related distress, interpersonal distress, physician-related distress, and emotional burden associated with living with diabetes. Items use Likert response categories reflecting frequency or intensity of distress experiences. Mean item scores were calculated according to established procedures; higher scores indicate greater diabetes-specific distress. The scale has demonstrated sensitivity to changes in diabetes management conditions, including adoption of monitoring technologies.^27^

A feasibility survey was specifically designed for this study by the research team. Throughout the study, participants were closely monitored for the presence of adverse events.

### Statistical analysis

Descriptive statistics were used to characterize the study population and main outcomes. Continuous variables were presented as median and interquartile range (IQR = P_25_ – P_75_) or mean and standard deviation (SD), as appropriate based on distribution. Categorical variables were summarized as absolute frequencies and percentages (n, %). Standardized scale and questionnaire scores (SUS, GMSS, patient and caregiver DDS, acceptability, and feasibility) were evaluated at both total and subscale levels using medians and IQRs. Additionally, scale responses were converted into predefined clinical or interpretive categories (e.g., satisfaction levels, distress severity thresholds, and Likert agreement categories) and reported as proportions (n, %). The data of participants who withdrew from the study was included up to the point of withdrawal in the analysis. All data analyses were performed using R statistical software (version 4.5.1; R Core Team, 2025).

## Results

### Baseline characteristics

A total of 40 participants were enrolled in the study between the 11^th^ of November 2025 and the 6^th^ of January 2026 after screening 95 participants for eligibility. One adult participant withdrew from the study midway after relocating out of the country, and one caregiver opted to withdraw their child due to frequent sensor detachment and inability to closely supervise her child. Their data up to the point of withdrawal was included in the analysis (baseline and CGM data) and they did not take part in the endline study surveys. The median age of participants was 13.0 (IQR, 9.3-17.5) years with majority (n = 19, 48%) being adolescents (online supplemental table 2). There were more females (n = 25, 63%) than males. Among caregivers, 93% (n = 28) were biological parents of these children. Their median age was 38 (IQR, 32-42) years with majority being women (n = 22, 73%).

### Usability

Overall, the median usability score was 98.8 (IQR, 92.5-100.0). Most participants (n = 36, 94.7%) reported *excellent* usability (scores > 80.3), while the remaining two participants (5.3%) achieved scores indicating *good* usability (≥ 70) (Figure 1).

At the item level, responses across usability domains were highly favorable (online supplemental figure 1). For positively worded statements, such as system ease of use, frequency of intended use, functional integration, confidence and quick learnability, participants demonstrated consistently high levels of agreement (95-97% “*Strongly Agree*”; 100% total agreement). Conversely, participants rejected negatively worded items: 100% disagreed that the system was unnecessarily complex or cumbersome to use (95% “*Strongly Disagree*”, 5% “*Disagree*”), and 95% disagreed (84% “*Strongly Disagree*”) with needing technical support. Regarding system consistency, 68% strongly disagreed that there was “*too much inconsistency”, while* 16% agreed and 5% remained neutral.

### Acceptability

Participant acceptability of CGM was exceptionally high across all evaluated domains (Figure 2). Combined positive ratings (“*very positive*” or “*positive*”) reached 100% for “general acceptability”, “affective attitude”, “self-efficacy”, “intervention coherence”, and “ethical consequences”. Specifically, “general acceptability” and “affective attitude” achieved the highest proportion of “*very positive*” responses (97% each). “Opportunity cost” and “perceived burden” also demonstrated near-unanimous positive endorsement (98%), indicating minimal lifestyle trade-offs and low daily effort. Finally, “perceived effectiveness” was rated favorably by 94% of participants (89% “*very positive*”), with 5% remaining neutral.

### Glucose monitoring satisfaction

Overall glucose monitoring satisfaction improved following the intervention (Figure 3). The mean total GMSS score increased from 3.75 ± 0.81 at baseline (n = 16) to 4.58 ±

0.42 at endline (n = 15). Individual trajectories showed that most participants improved, while two experienced a decline and one showed no change. Correspondingly, the proportion of participants with high satisfaction increased from 38% (n = 6/16) at baseline to 80% (n = 12/15) at endline, with low satisfaction dropping from 13% (n = 2/16) to 0% (online supplemental table 3).

At the subscale level, positive trends were observed in all domains (Figure 3). User strain was markedly reduced, as evidenced by lower mean scores in “emotional burden” (2.19 ± 0.90 vs. 1.22 ± 0.42) and “behavioral burden” (2.14 ± 1.06 vs. 1.32 ± 0.43). Concurrently, mean scores for “openness” (3.55 ± 0.92 versus 4.43 ± 0.59) and “trust” (3.79 ± 1.16 versus 4.38 ± 0.85) increased, despite minor individual variation.

### Diabetes distress

#### Adolescents (aged 15 years and above) and adults

Overall diabetes distress was low at baseline and decreased further following the intervention, with median total scores changing from 1.54 (IQR, 1.00-1.96) to 1.36 (IQR, 1.00-1.82) (Figure 4). At baseline, two participants (2/15, 13.3%) reported high total diabetes distress (score > 3), whereas no participants (0/14, 0%) exhibited high distress at endline (online supplemental table 4). At the subscale level, median distress scores declined across most domains. The only exceptions were “physician distress”, which remained unchanged at a minimum floor median of 1.00, and “hypoglycemia distress”, which showed a slight increase in median score from 1.00 (IQR, 1.00 – 1.75) to 1.25 (IQR, 1.00-1.50).

### Caregivers DDS

Caregiver diabetes distress decreased overall following the intervention (online supplemental table 5). The median total score reduced from 2.05 (IQR, 2.00–2.75; moderate distress) at baseline to 1.90 (IQR, 1.00–2.60; low distress) at endline. High total distress (score > 3) was reported by 13.3% of caregivers (n = 4/30) at baseline, decreasing to 3.5% (n = 1/29) at endline. As illustrated in Figure 5, median distress scores declined in “teen management” (2.63 to 2.25) and “parent/teen relationship distress” (1.94 to 1.75), remained stable for “health care team distress” (median 1.00), and slightly increased for “personal distress” (2.17 to 2.33).

### Feasibility

Most participants achieved consistently high wear time of ≥70-100% (Supplementary figure 2). The median CGM active wear time across the enrolled population was 89% (IQR, 75-92%), with 80.0% (n = 32/40) of participants maintaining a total wear time exceeding 70%. Nevertheless, intermittent gaps in wear time were observed across participants, indicating episodic interruptions in CGM use.

Daily scanning frequency remained relatively stable throughout the study, typically ranging between 6 and 10 scans per day (online supplemental figure 3). However, notable inter-individual variability was observed, with occasionally daily spikes exceeding 60–80 scans per day in some individuals (online supplemental figure 4). Importantly, there was no clear downward trend across the three-month period (M1 to M3), demonstrating sustained participant engagement over time.

All participants described the CGM application process as “very easy” (n = 32, 84.2%) or “easy” (n = 6, 15.8%), with 97.4% (n = 37) participants reporting that they never experienced difficulties with CGM placement during the study period (online supplemental table 6). Most participants (n = 32, 84.2%) reported that the CGM device never fell off during the study period, while 13.2% (n = 5) reported that it had fallen off once or twice, and 2.6% (n = 1) reported three to four detachments. Following sensor removal, 63.2% (n = 24) reapplied a new sensor immediately without delay, while 36.8% (n = 14) reported that they had delayed a few times. Skin irritation or allergic reactions were experienced by 7.9% (n = 3) of participants.

## Discussion

In this prospective review of 40 youth with T1D at Kenyatta National Hospital in Nairobi, there was high usability, acceptability, and feasibility of CGM, as well as improved glucose monitoring satisfaction, and reduced diabetes-related distress among both adolescents and caregivers.

There was a high median SUS score of 98.8 in our study, which was comparable to that by Psavko et al., who reported a score of 92.6 among adults with diabetes, and diabetes care and education specialists.^28^ In contrast, an online survey of 281 Italian patients found a lower average score of 66.^29^ The high scores in the current study indicate a positive user experience among this population. Similarly, in a prospective randomized trial that assessed CGM versus SMBG only in 158 adults with poorly controlled T1D, Polonsky et al. found that satisfaction with CGM was high with perceived benefits noted as very common, and perceived hassles reported as relatively rare.^30^ Our findings suggest that usability would not be a hindrance to CGM adoption in this population. There was high acceptability in all domains in the present study, which is important for sustained adherence. Most participants felt confident in using CGM effectively and reported low perceived burden suggesting that it could be integrated into their daily life without disruption. Similarly, qualitative results from a clinical trial conducted in the region showed that CGM was feasible and acceptable within the South Africa healthcare context.^31^

Our study showed an improvement in glucose monitoring satisfaction. This finding is consistent with a systematic review and meta-analysis which found that, out of 43 studies assessing CGM, 88% reported improved patient satisfaction, and all the studies reported increased hypoglycemia awareness.^32^ Patient satisfaction may influence self-management behaviours and sustained device use, affecting short and long-term clinical outcomes. In the current study, there were improvements in all GMSS domains. The reduced “behavioural burden” was likely due to the reduced effort required for glucose monitoring with CGM, compared to SMBG. Similarly, the reduced “emotional burden” may be due to improved visibility of glucose trends, thus increasing the user’s sense of control and reducing anxiety. The increase in “openness” scores indicated that with CGM, participants felt less restricted by diabetes and were more open to new experiences in life. The increased “trust” in glucose monitoring data was important as it influences the willingness of the user to respond to glucose data. Of note, a few participants experienced a reduction in glucose monitoring satisfaction. Similar findings have been reported in systematic reviews that have found that not all patients experience psychosocial benefits.^33^ These varying responses may be due to alarm fatigue, technical challenges, anxiety due to increased data visibility, and body image issues, highlighting the importance of continued education and psychosocial support.

In our study, there was a modest reduction in diabetes distress among adolescents and caregivers after the intervention, a finding that is similar to that reported in other studies.^30,34,35^ One meta-analysis found that out of 43 studies, 56% reported significant reductions in diabetes distress.^32^ These findings suggest use of CGM may improve confidence in glucose management. The increase in hypoglycemia-related distress was, however, of concern. In contrast, Polonsky et al. noted that participants using CGM had better hypoglycemic confidence than those using SMBG, a finding that is supported by others.^30,35^ Our study finding may be due to anxiety and fatigue resulting from hypoglycemia alerts, highlighting the need for ongoing psychological support. The reduced overall distress among caregivers in the current study is in keeping with other studies, a finding that is indicative of the psychosocial benefits of CGM.^36^ The improvements observed in our study in “teen management” and “parent–teen relationship distress” were likely due to less parent-teen conflict. The slight increase in “personal distress” among caregivers, however, may possibly have been due to increased vigilance due to better glucose data visibility.

All the participants in the present study described the application process as either easy or very easy, indicating that CGM insertion procedures are user-friendly.^37^ CGM scanning among participants in our study did not diminish over time. However, there were very high scanning frequencies among some participants, which may have been due to increased vigilance. Meguro et al. in Japan found that psychological factors such as behaviour towards hypoglycemia contributed to scanning frequency. ^38^ This highlights the importance of individualized education. Although the current study was carried out over a short period of three months, and scanning frequency might decrease over time due to fatigue, the sustained engagement observed supports the feasibility of CGM use in this population.

The median sensor wear time in our study was high at 89%. Our findings are similar to those of Baker et al, who in a prospective study of a cohort of 50 patients with T1D in Rwanda, found that participants used the CGM more than 80% of the time.^20^ Eighty-four percent of participants in the present study reported that the CGM sensor never detached during the study period. The detachment rate of 16% was, however, higher than reported in a study among 42 children in India.^17^ Physical activity, limited body surface area in smaller children and ambient temperature and humidity can influence sensor adhesion.^38^ Adhesive products and external wraps, as well as patient education, can help reduce detachment rates.^39^

In this study, 63% of participants did not report delays in sensor reapplication, while the rest (37%) experienced delays, resulting in data gaps. Among the seven participants, the most commonly cited reason for delayed reapplication was the lack of an extra sensor. In a meta-analysis of randomized controlled trials assessing glycemic control in T1D among those using CGM versus those using SMBG, it was found that CGM was associated with significant improvements in HbA1c among those who used the sensors most frequently.^40^ In the current study, only eight percent of participants reported experiencing skin irritation or allergic skin responses, an incidence that was lower than reported in other studies.^17,41^ Our finding may possibly due to under-reporting of mild symptoms, low prevalence of atopy, and genetic or immunologic differences. The low rate supports long-term feasibility of CGM adoption in this population.

The high usability, acceptability, and feasibility of CGM in the current study has important implications for policy and clinical practice, indicating that these three outcomes measures are unlikely to be a barrier to its adoption by users. These findings support its wider implementation among children, adolescents, and young adults with T1D in low-resource settings as part of standard of care to maximize clinical benefits. Finally, health systems that support sustained access to devices and continuous training of health care providers are essential to ensure long-term success of CGM implementation.

## Strengths and limitations

The comprehensive evaluation of CGM across usability, acceptability, and feasibility outcomes, with the consistency of findings, was the main strength of this study. The main limitation was that the short follow-up period does not capture changes in users’ perceptions or adherence during long-term CGM use. There may also have been response bias due to the excitement of a new intervention. This study, however, forms the baseline for future studies.

In conclusion, there was high usability, acceptability, and feasibility of CGM among youth with T1D, with sustained engagement, improved glucose monitoring satisfaction, and modest reductions in diabetes distress. This supports the integration of CGM into routine diabetes management, and its broader implementation in low-resource settings. Further research should explore long-term usage patterns and psychological responses in this setting.

## Footnotes

### Contributors

Study conception and design: EMC, YK, CH, PA, and LM. Training: PA, LM, AK, YK, JK, and EMC. Data acquisition: PA, LM, WM, JK, and AK. Funding acquisition/resources: CH. Data analysis and visualization: NB. Data analysis and interpretation: PA, LM, EMC, NB, YK, JK, CH, and AK. Original manuscript draft preparation: PA. All authors contributed to the final manuscript.

### Funding

This work was supported by the Foundation for Innovative New Diagnostics (FIND), with a grant from The Leona M. and Harry B. Helmsley Charitable Trust.

### Conflict of Interest

The authors have no conflicts of interest to declare that are relevant to the content of this article.

### Data Availability Statement

The data used to support the findings of this study are restricted by the Kenyatta National Hospital-University of Nairobi Ethics Review Committee, to protect patient privacy. It may be released upon application to the ethical board, who can be contacted at.

### Ethics statements

The study was conducted after obtaining written approval from the Kenyatta National Hospital-University of Nairobi Ethics and Research Committee (Study Number P291/03/2025, approval date 17^th^ July 2025, amendment approval date 27^th^ November 2025) and the National Commission for Science, Technology and Innovation. Approval to conduct the study on human participants was obtained from the Pharmacy and Poisons Board as per the laws of Kenya; approval number ECCT/25/08/07.

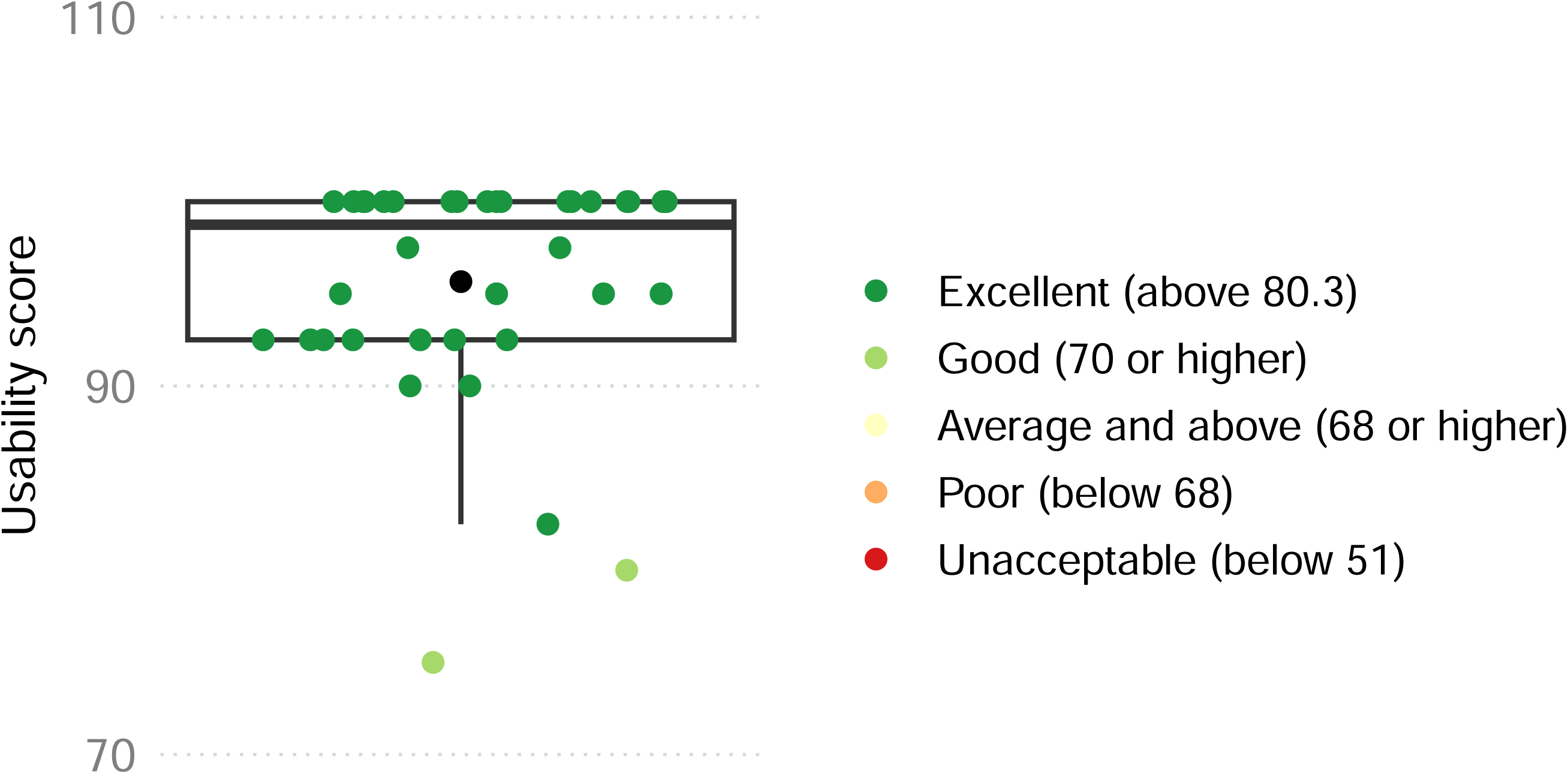

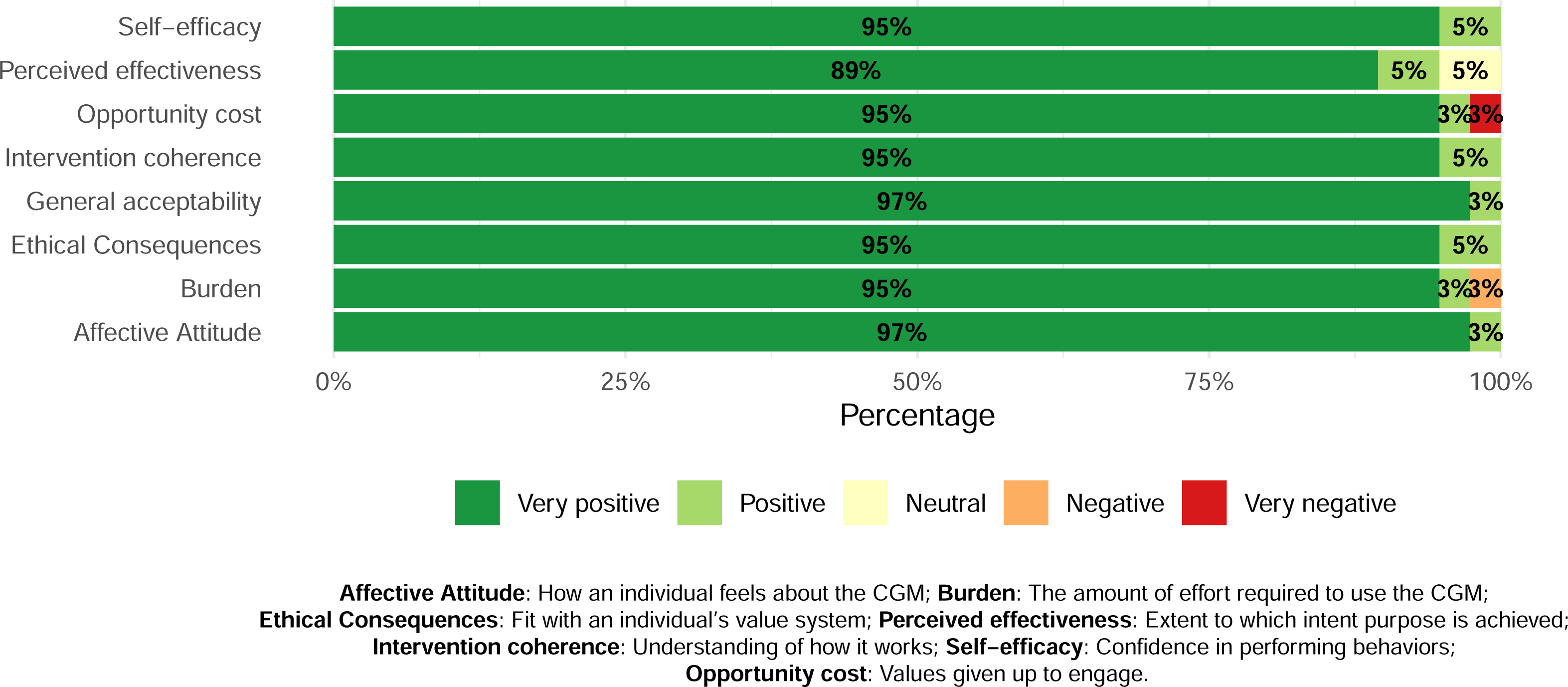

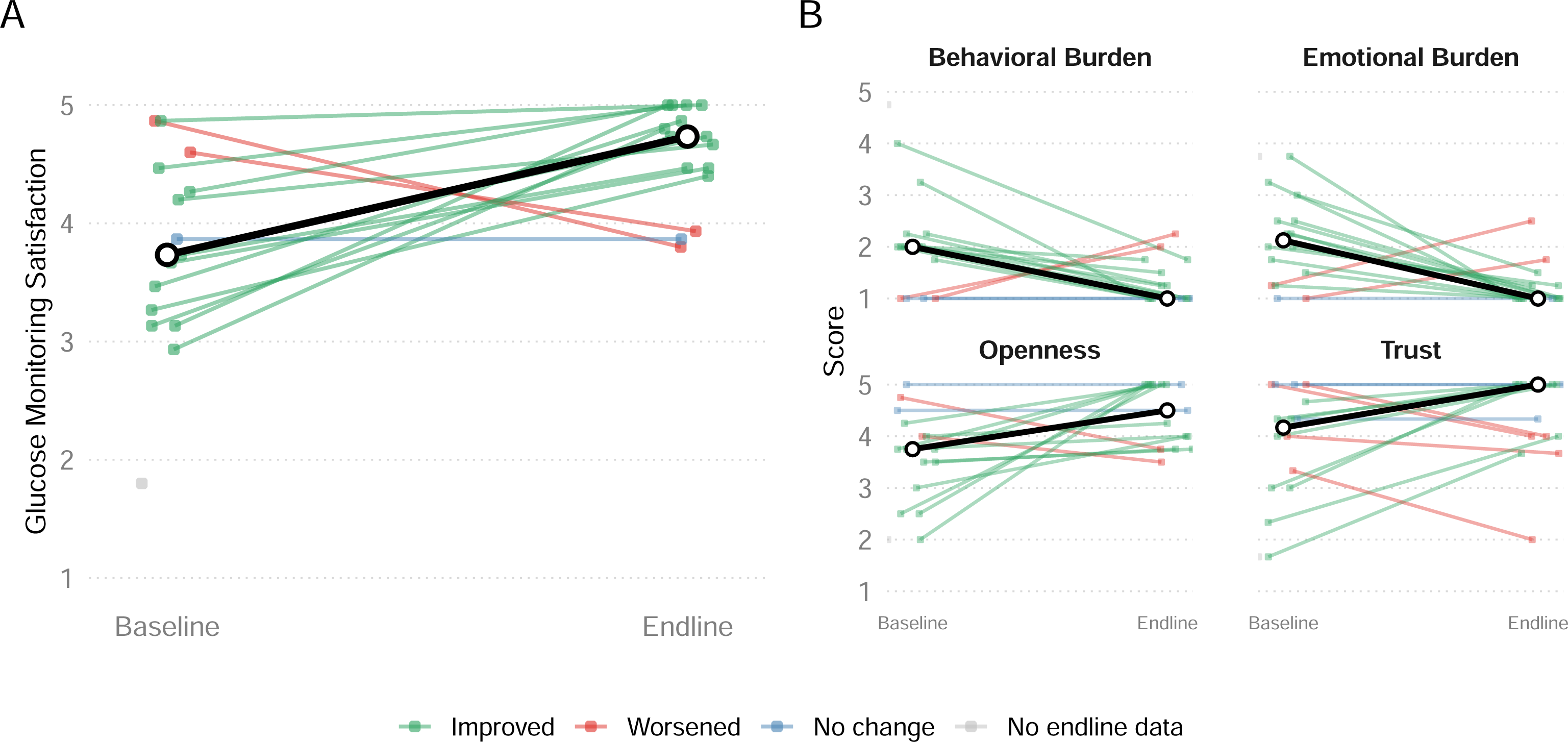

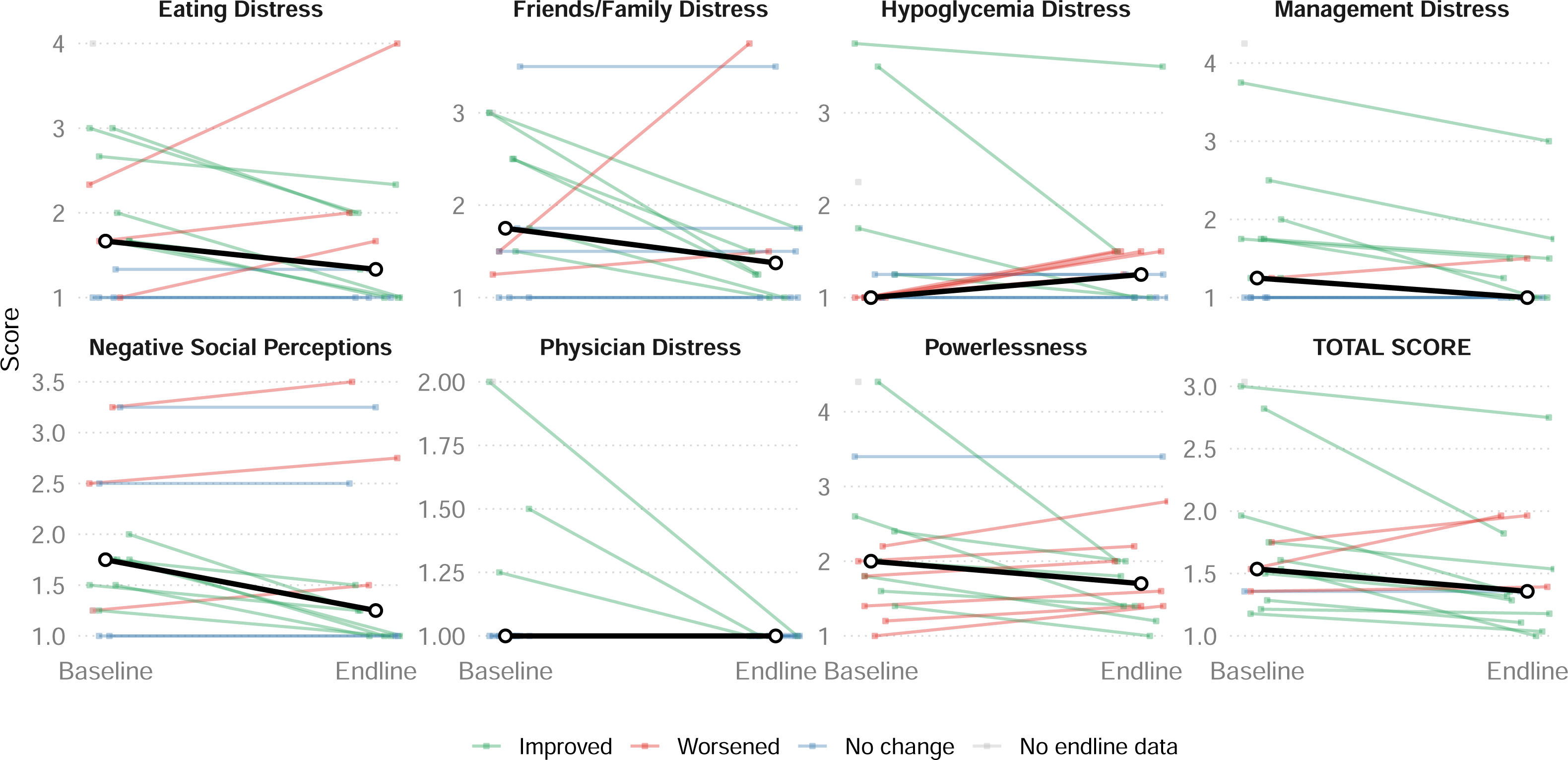

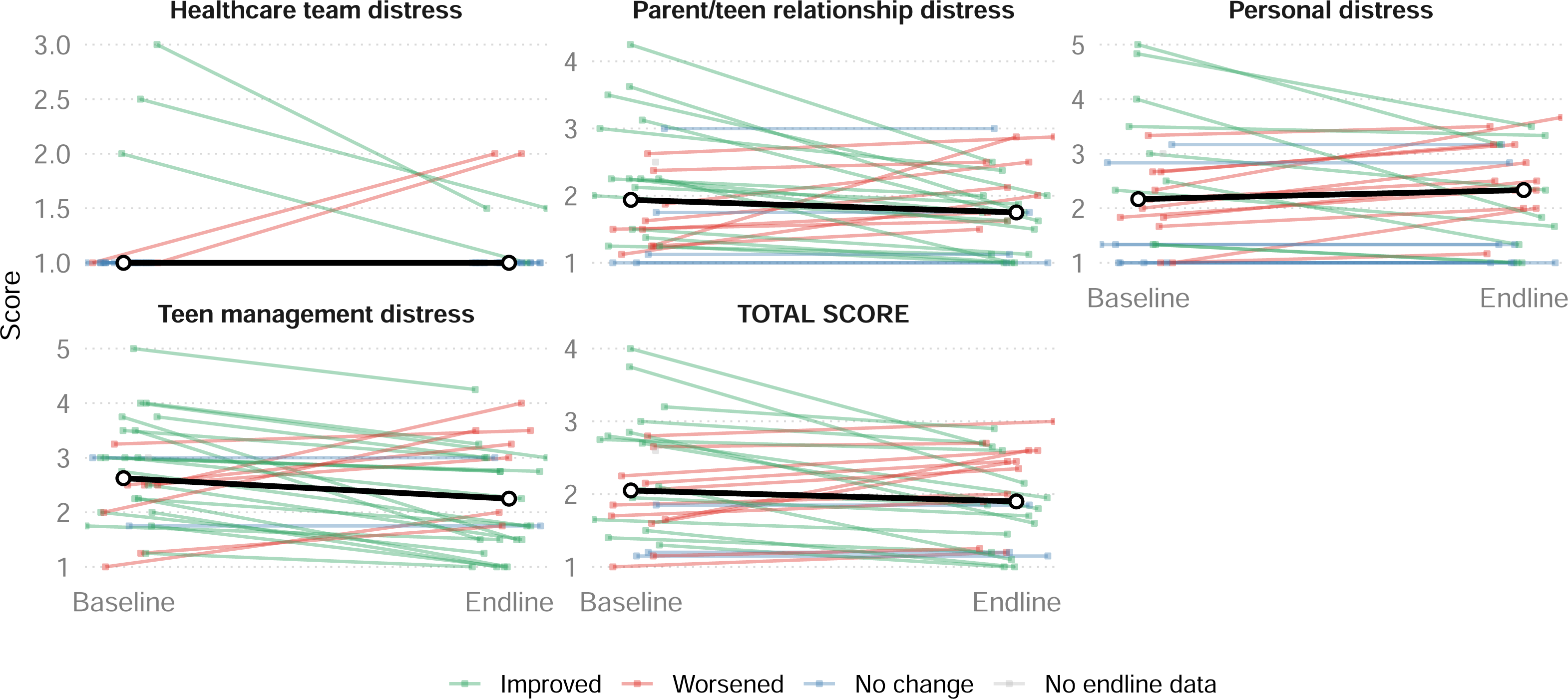

## Notes

### Competing Interest Statement

The authors have declared no competing interest.

### Author Declarations

Kenyatta National Hospital-University of Nairobi Ethics and Research Committee of University of Nairobi gave ethical approval for this work.

