## Supplemental Table 1 for "Usability, acceptability and feasibility of continuous glucose monitoring among children and adolescents with type 1 diabetes in Kenya"

### SUPPLEMENTARY MATERIALS

#### Supplementary Table 1

Schedule of activities.

| | **Visit 1: Baseline** | **Visit 2** | **Visit 3** | **Visit 4** | **Visit 5: Endline** |
|---|---|---|---|---|---|
| Study procedures | Enrolment and informed consent | | | | Completion log |
| Doctors' visit | Standard of care visit | | | Standard of care visit | Standard of care visit |
| CGM procedures | | CGM device application and distribution of CGM devices | CGM device removal.<br>New CGM device application.<br>CGM data was downloaded and the doctor explained it to the participant. | CGM data was downloaded and the doctor explained it to the participant. | CGM data was downloaded and the doctor explained it to the participant. |
| Diabetes education | Diabetes education session on CGM use | | Education on CGM removal procedure | Diabetes education based on the CGM data | Diabetes education based on the CGM data |
| Completion of surveys/ CRFs | 1. Sociodemographic survey<br>2. DDS (and/or parent DDS)<br>3. GMSS (referred to SMBG) | | | | 1. Usability survey<br>2. DDS (and/or parent DDS)<br>3. GMSS (referred to CGM)<br>4. Acceptability |
| Safety | From baseline to endline: monitoring for SAEs | | | | |

CGM: Continuous Glucose Monitoring; DDS: Diabetes Distress Scale; GMSS: Glucose Monitoring Satisfaction Survey; SAE: Serious Adverse Event. *DDS was responded to by participants 15 years old and older. Parent DDS was responded to by all caregivers of minor participants (<18 years old). **GMSS was responded to by participants 14 years old and older.
