## Supplemental Table 2 for "Usability, acceptability and feasibility of continuous glucose monitoring among children and adolescents with type 1 diabetes in Kenya"

**Supplementary Table 2**

Baseline Characteristics.

| | **N = 40[1]** |
|---|---|
| Sex: Females | 25 (63%) |
| Age (years) | 13.0 (9.3, 17.5) |
| Age | |
| Paediatric (4 to under 10 years old) | 11 (28%) |
| Adolescent (10 to under 18 years old) | 19 (48%) |
| Adult (18 - 25 years old) | 10 (25%) |
| Highest level of education completed | |
| Primary | 25 (63%) |
| Secondary/A level | 10 (25%) |
| College/Middle level | 1 (2.5%) |
| University | 4 (10%) |
| Employment status | |
| Employed full time | 2 (5.0%) |
| Employed part time | 1 (2.5%) |
| Student | 3 (7.5%) |
| None | 4 (10.0%) |
| Not Applicable (for children/adolescents) | 30 (75.0%) |
| Marital status | |
| Single | 10 (25.0%) |
| Not Applicable (for children/adolescents) | 30 (75.0%) |
| Household income, in KES | |
| < = 19, 999 | 2 (5.0%) |
| 20,000 - 39,999 | 1 (2.5%) |
| Don’t Know | 7 (18.0%) |
| Not Applicable (for children /adolescents below 18 years) | 30 (75.0%) |
| History of comorbidities | 5 (12.5%) |
| HbA1c (%) at enrollment | 10.6 (9.0, 12.0) |
| Years living with T1D | 5.0 (3.0, 6.5) |

| | |
|---|---|
| Average times checking blood glucose levels per day | |
| 0 | 0 (0%) |
| 1-3 | 21 (52.5%) |
| 4-6 | 19 (47.5%) |
| Number of hypoglycemic episodes per week | |
| 0 | 7 (17.9%) |
| 1-2 | 19 (48.7%) |
| More than 2 | 13 (33.3%) |
| Unknown | 1 |
| Number of DKA within the last 12 months | |
| 0 | 23 (57.5%) |
| 1 | 10 (25.0%) |
| More than 1 | 7 (17.5%) |
| Combinations of type(s) of insulin currently used | |
| Rapid-acting insulin and Long-acting insulin | 7 (17.5%) |
| Short-acting insulin and Long-acting insulin | 33 (82.5%) |
| History using CGM device | 4 (10.0%) |
| History using CGM device | |
| Freestyle Libre | 3 (7.5%) |
| Other | 2 (5.0%) |
| **Caregiver's characteristics (N = 30)** | |
| Relationship to the child with Type 1 diabetes | |
| Biological Father/Mother | 28 (93.3%) |
| Uncle/Aunt | 2 (6.7%) |
| Caregiver's age | 38 (32, 42) |
| Gender: Women | 22 (73.3%) |
| Caregiver's education | |
| Primary | 6 (20.0%) |
| Post-primary/Vocational | 2 (6.7%) |
| Secondary/A level | 7 (23.3%) |
| College/Middle level | 12 (40.0%) |
| University | 3 (10.0%) |

| | |
|---|---|
| Caregiver's employment status | |
| Employed full time | 6 (20.0%) |
| Employed part time | 8 (26.7%) |
| Self-employed/Freelance | 11 (36.7%) |
| Homemaker | 5 (16.7%) |
| Caregiver's marital status | |
| Single | 5 (16.7%) |
| Married/Civil Union | 20 (66.7%) |
| Divorced/Separated | 2 (6.7%) |
| Widowed | 3 (10.0%) |
| Household income (in KES) | |
| < = 19, 999 | 12 (40.0%) |
| 20,000 - 39,999 | 7 (23.3%) |
| 40,000 - 59,000 | 5 (16.7%) |
| 60,000 - 79,000 | 3 (10.0%) |
| 80,000-99,999 | 1 (3.3%) |
| >= 100,000 | 2 (6.7%) |

[1]n (%); Median (Q1, Q3)
