## Supplementary figures and images for "Usability, acceptability and feasibility of continuous glucose monitoring among children and adolescents with type 1 diabetes in Kenya"

### Supplemental Figure 1

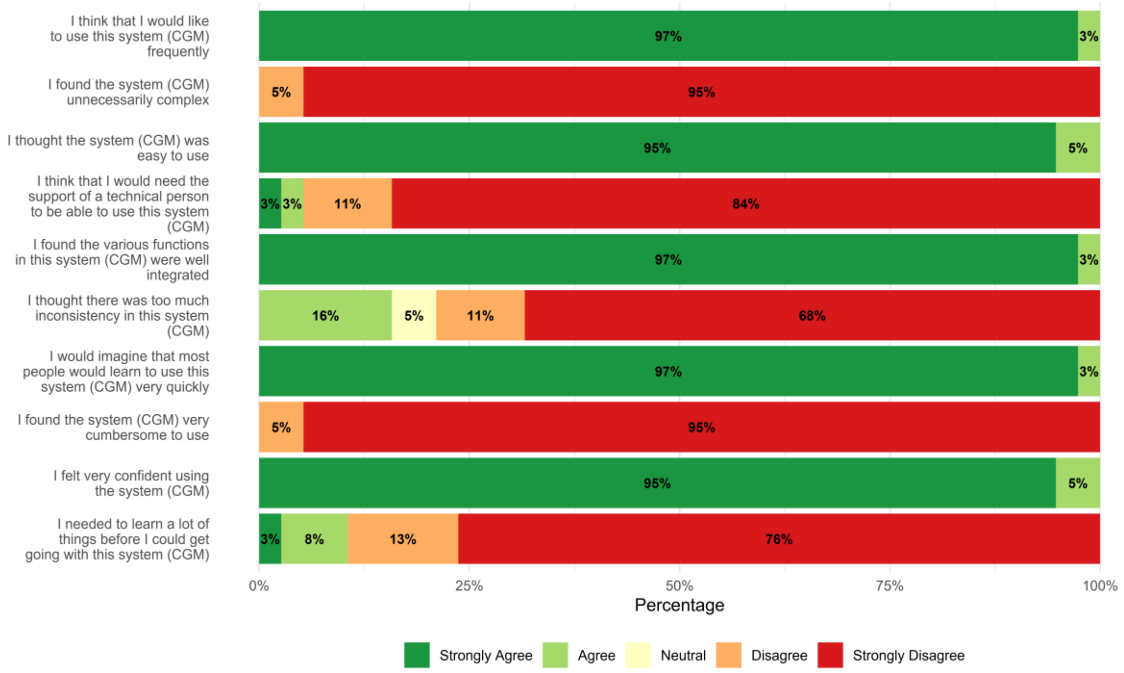


#### Supplementary Figure 1

Usability domains at endline.
