## Supplemental Table 3 for "Usability, acceptability and feasibility of continuous glucose monitoring among children and adolescents with type 1 diabetes in Kenya"

**Supplementary Table 3**

Glucose monitoring satisfaction survey.

| | Baseline<br>N = 16 | Endline<br>N = 15 |
|---|---|---|
| **Total score** | | |
| *Mean (SD)* | 3.75 (0.81) | 4.58 (0.42) |
| **GMS** | | |
| *Low satisfaction* | 2 (12.5%) | 0 (0.0%) |
| *Moderate satisfaction* | 8 (50.0%) | 3 (20.0%) |
| *High satisfaction* | 6 (37.5%) | 12 (80.0%) |
| **Openness subscale** | | |
| *Mean (SD)* | 3.55 (0.92) | 4.43 (0.59) |
| **Emotional Burden subscale** | | |
| *Mean (SD)* | 2.19 (0.90) | 1.22 (0.42) |
| **Behavioral Burden subscale** | | |
| *Mean (SD)* | 2.14 (1.06) | 1.32 (0.43) |
| **Trust subscale** | | |
| *Mean (SD)* | 3.79 (1.16) | 4.38 (0.85) |
