## Supplemental Table 4 for "Usability, acceptability and feasibility of continuous glucose monitoring among children and adolescents with type 1 diabetes in Kenya"

**Supplementary Table 4**

Participant's Diabetes Distress Score.

| | Baseline, N = 15 | | | Endline, N = 14 | | |
|---|---|---|---|---|---|---|
| | Median (Q1, Q3) | Moderate distress (score 2-2.9) | High distress (score >3) | Median (Q1, Q3) | Moderate distress (score 2-2.9) | High distress (score >3) |
| **Total Participant DDS** | **1.54 (1.00-1.96)** | **1 (6.7%)** | **2 (13.3%)** | **1.36 (1.00-1.82)** | **1 (7.1%)** | **0 (0.0%)** |
| Powerlessness | 2.00 (1.00-2.60) | 5 (33.3%) | 3 (20.0%) | 1.70 (1.00-2.00) | 5 (35.7%) | 1 (7.1%) |
| Management Distress | 1.25 (1.00-2.00) | 2 (13.3%) | 2 (13.3%) | 1.00 (1.00-1.50) | 0 (0.0%) | 1 (7.1%) |
| Hypoglycemia Distress | 1.00 (1.00 - 1.75) | 1 (6.7%) | 2 (13.3%) | 1.25 (1.00-1.50) | 0 (0.0%) | 1 (7.1%) |
| Negative Social Perceptions | 1.75 (1.00 - 2.50) | 3 (20.0%) | 2 (13.3%) | 1.25 (1.00-1.50) | 2 (14.3%) | 2 (14.3%) |
| Eating Distress | 1.67 (1.00-2.67) | 3 (20.0%) | 3 (20.0%) | 1.33 (1.00-2.00) | 4 (28.6%) | 1 (7.1%) |
| Physician Distress | 1.00 (1.00-1.25) | 2 (13.3%) | 0 (0.0%) | 1.00 (1.00-1.00) | 0 (0.0%) | 0 (0.0%) |
| Friend/Family Distress | 1.75 (1.00-3.00) | 2 (13.3%) | 4 (26.7%) | 1.38 (1.00-1.75) | 0 (0.0%) | 2 (14.3%) |
