## Supplemental Table 5 for "Usability, acceptability and feasibility of continuous glucose monitoring among children and adolescents with type 1 diabetes in Kenya"

**Supplementary Table 5**

Caregiver Diabetes Distress Score.

| | Baseline, N = 30 | | | Endline, N = 29 | | |
|---|---|---|---|---|---|---|
| | **Median (Q1, Q3)** | **Moderate distress (score 2-2.9)** | **High distress (score >3)** | **Median (Q1, Q3)** | **Moderate distress (score 2-2.9)** | **High distress (score >3)** |
| **Total Caregiver DDS** | **2.05 (2.00-2.75)** | **12 (40.0%)** | **4 (13.33%)** | **1.90 (1.00-2.60)** | **12 (41.4%)** | **1 (3.5%)** |
| Personal Distress | 2.17 (1.00-3.00) | 9 (30.0%) | 8 (26.7%) | 2.33 (1.00-3.17) | 9 (31.0%) | 8 (27.6%) |
| Teen Management Distress | 2.63 (2.00-3.25) | 10 (33.3%) | 14 (46.7%) | 2.25 (2.00-3.00) | 5 (17.2%) | 11 (37.9%) |
| Prent/Teen Relationship Distress | 1.94 (1.00 - 2.50) | 9 (30.0%) | 6 (20.0%) | 1.75 (1.00-2.13) | 10 (34.5%) | 1 (3.5%) |
| Health Care Team Distress | 1.00 (1.00 - 1.00) | 2 (6.7%) | 1 (3.3%) | 1.00 (1.00-1.00) | 2 (6.9%) | 0 (0.0%) |
