## Supplemental Figure 2 for "Usability, acceptability and feasibility of continuous glucose monitoring among children and adolescents with type 1 diabetes in Kenya"

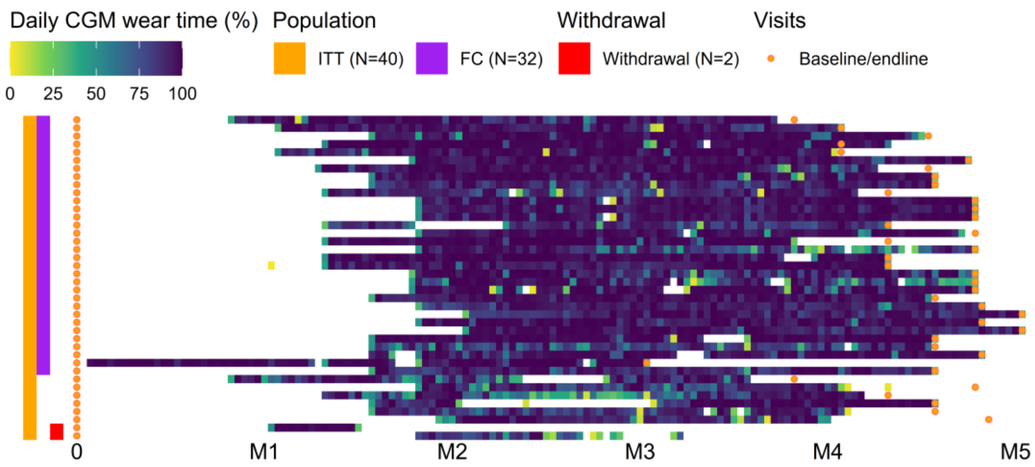


**Supplementary Figure 2**

Heatmap of daily CGM wear time (%) among 40 participants. Each row represents an individual participant, and each column represents a study day from baseline (0) through months 1 to 5 (M1–M5). Color intensity reflects the proportion of daily wear time, ranging from low (0%) to high (100%). Orange and purple indicators denote the intention-to-treat (ITT, N=40) and fully compliant (FC, N=32) (sensor wear time above 70%) populations, respectively. Red markers indicate participants who withdrew (N=2), and orange dots represent baseline and endline study visits. M: Month.
