## Supplemental Figure 3 for "Usability, acceptability and feasibility of continuous glucose monitoring among children and adolescents with type 1 diabetes in Kenya"

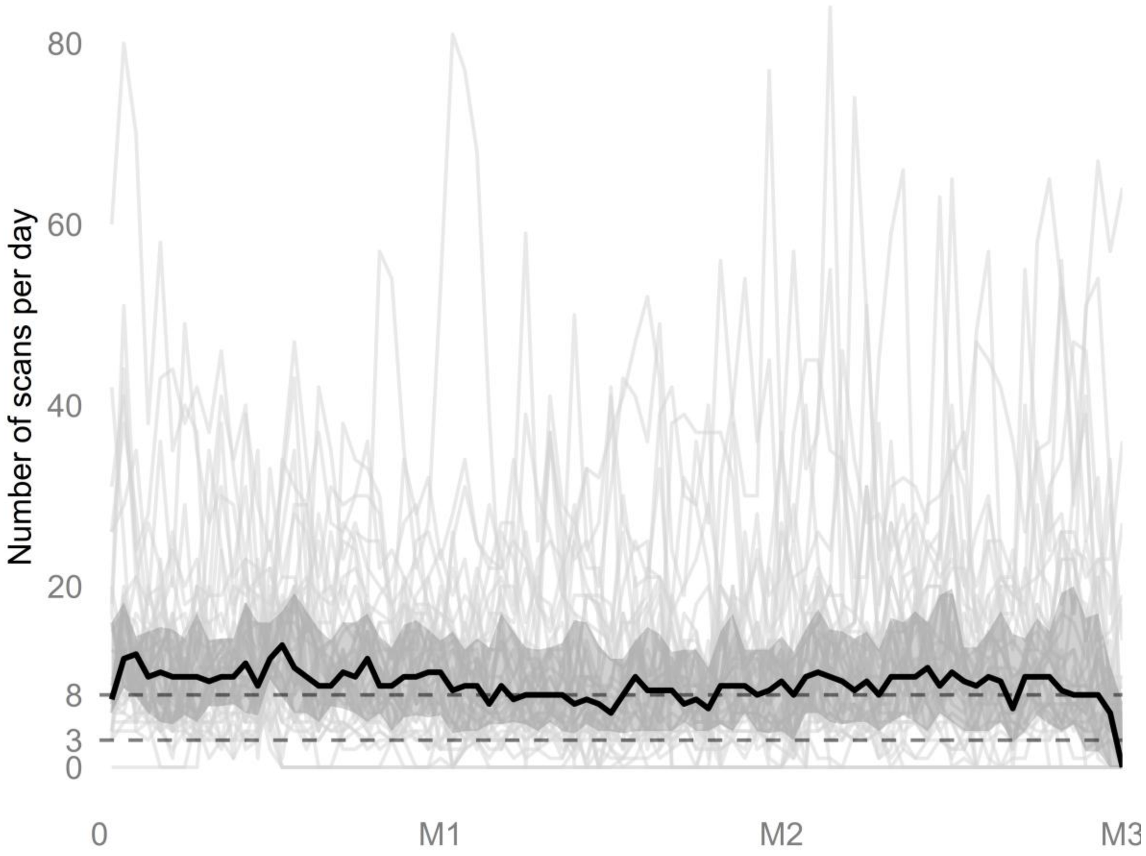


**Supplementary Figure 3**

Daily CGM scanning frequency over 3 months. The black line represents the median; the shaded ribbon represents the IQR (Q1-Q3), and light gray lines show individual participant data. M: month.
