## Supplemental Figure 4 for "Usability, acceptability and feasibility of continuous glucose monitoring among children and adolescents with type 1 diabetes in Kenya"

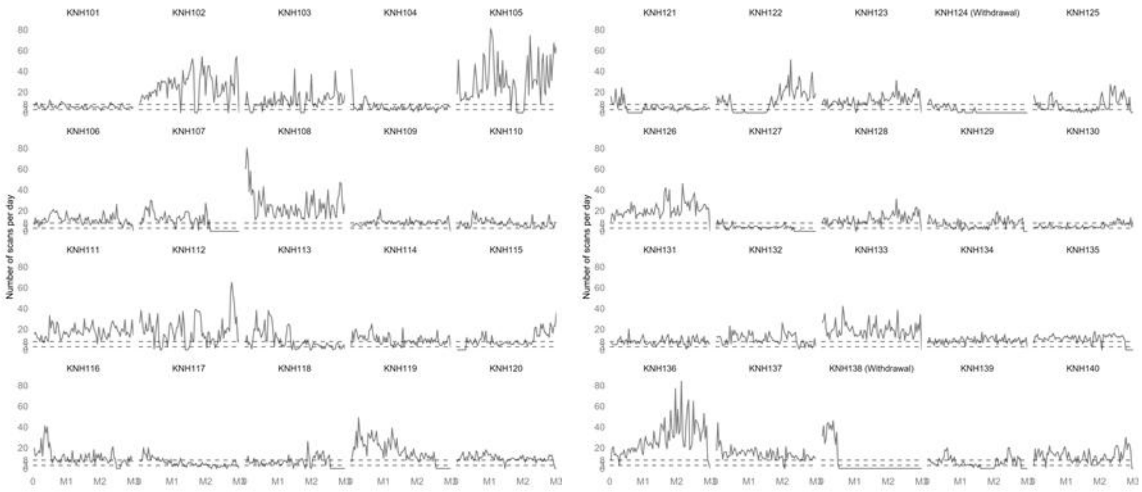


**Supplementary Figure 4**

Daily CGM scanning frequency over 3 months. Each plot shows individual participant data.
