## Supplemental Table 6 for "Usability, acceptability and feasibility of continuous glucose monitoring among children and adolescents with type 1 diabetes in Kenya"

**Supplementary Table 6**

Feasibility items at endline.

| | **N = 38**[1] |
|---|---|
| How easy or difficult was it to apply the CGM? | |
| Very easy | 32 (84.2%) |
| Easy | 6 (15.8%) |
| Neutral | 0 (0.0%) |
| Somewhat difficult | 0 (0.0%) |
| Very difficult | 0 (0.0%) |
| How frequently did you experience difficulties with insertion or placement of the CGM device? | |
| Never (0) | 37 (97.4%) |
| Rarely (1-2 times per month) | 1 (2.6%) |
| Occasionally (3-4 times per month) | 0 (0.0%) |
| Frequently (5-6 times per month) | 0 (0.0%) |
| Very frequently (7 or more times per month) | 0 (0.0%) |
| How many times did your CGM fall off in the past 3 months? | |
| Never | 32 (84.2%) |
| Few times (1-2) | 5 (13.2%) |
| Sometimes (3-4) | 1 (2.6%) |
| Frequently (5-6) | 0 (0.0%) |
| Very frequently (>6) | 0 (0.0%) |

| | |
|---|---|
| Were there times when you delayed reapplying the CGM after removal? | |
| Never | 24 (63.2%) |
| Few times (1-2) | 14 (36.8%) |
| Sometimes (3-4) | 0 (0.0%) |
| Frequently (5-6) | 0 (0.0%) |
| Very frequently (>6) | 0 (0.0%) |
| How frequently did you experience skin irritation or allergic reactions wearing the CGM? | |
| Never (0) | 35 (92.1%) |
| Rarely (1-2 times per month) | 3 (7.9%) |
| Occasionally (3-4 times per month) | 0 (0.0%) |
| Frequently (5-6 times per month) | 0 (0.0%) |
| Very frequently (7 or more times per month) | 0 (0.0%) |
